## Supplemental Appendix 1 for "Estimating time-varying cholera transmission and oral cholera vaccine effectiveness in Haiti and Cameroon, 2021-2023" (from another version of this preprint: version 1, 2024-06-13)

### Appendix 1: Supplemental methods and results for **"Estimating time-varying transmission and oral cholera vaccine effectiveness in Haiti and Cameroon, 2021-2023"**

Erin N. Hulland[1,2], Marie-Laure Charpignon[2,3], Ghinwa Y. El Hayek[2], Lihong Zhao[2,4], Angel N. Desai[2,5], Maimuna S. Majumder[1,2]

[1] Computational Health Informatics Program, Boston Children's Hospital & Harvard Medical School, Boston, MA, United States

[2] Comp Epi Dispersed Volunteer Research Network, Boston, MA, United States

[3] Institute for Data, Systems, and Society, Massachusetts Institute of Technology, Cambridge, MA, United States

[4] Department of Mathematics, Virginia Tech, Blacksburg, VA, United States

[5] Department of Internal Medicine, Division of Infectious Diseases, University of California, Davis Health Medical Center, Sacramento, CA, United States

**Disclaimer: This work is not yet finalized and will be continuously updated. We welcome feedback and suggestions on the analyses and interpretation and any meaningful additions.**

**June 11, 2024**

### Table of Contents

**Table of Contents** 1
**Section 1: Additional Tables** 2
Table 1: Descriptive and modeled statistics from the literature, Cameroon and Haiti 2
Table 2: Number and percent of vaccine effectiveness observations outside 0% or 100%, Cameroon and Haiti 3
**Section 2: Sensitivity Analyses** 4
Artificially changing OCV coverage to understand impact on vaccine effectiveness, Cameroon and Haiti 4
Figure 1: Impact of assumed OCV1 coverage on vaccine effectiveness 4
Exploring alternate Rt windows in EpiEstim, Cameroon and Haiti 4
Figure 2: Impact of length of time window on Rt 5
Exploring alternate SI parameters in EpiEstim, Cameroon and Haiti 5
Figure 3: Impact of changing the SI on Rt 6
Exploring alternate values of R0 on vaccine effectiveness, Cameroon and Haiti 6
Figure 4: Impact of changing R0 on OCV1 effectiveness 7
Exploring the vaccine effectiveness in Ouest department, Haiti 7
Figure 5: Focusing on Ouest department for vaccine effectiveness in Haiti 8
**Section 3: References** 9

### Section 1: Additional Tables

Table 1: Descriptive and modeled statistics from the literature, Cameroon and Haiti

| | **Cameroon** | **Haiti** |
|---:|---:|---:|
| **2017 % with access to improved water source[1]** | 73.95 (71.24-76.90) | 68.29 (66.32-70.19) |
| **2017 % with access to improved sanitation[1]** | 52.93 (51.05-55.11) | 51.50 (50.31-52.79) |
| **2017 under-5 mortality per 1,000 live births[2]** | 73.5 (60.5-87.0) | 59.3 (50.0-71.0) |
| **2019 one-dose MCV coverage[3]** | 67.8 (59.2-75.2) | 71.6 (65.1-77.4) |
| **% of children with diarrhea taking ORT[4]** | 19.91 (11.95-31.05) | 39.05 (29.17-49.41) |
| **2017 mean years education: female 15-49[5]** | 7.77 (6.72-8.79) | 6.42 (5.70-7.05) |
| **2017 difference in mean years education males to females 15-49 years[5]** | 0.66 (0.48-0.93) | 0.71 (0.66-0.88) |
| **2019 under-5 diarrhea prevalence[6]** | 43.17 (31.79-58.94) | 48.51 (44.42-52.28) |
| **2019 under-5 stunting prevalence[7]** | 33.07 (26.41-40.55) | 23.79 (18.91-29.41) |
| **2019 under-5 wasting prevalence[7]** | 6.30 (4.31-8.94) | 6.25 (3.21-10.41) |
| **2019 under-5 underweight prevalence[7]** | 12.77 (9.94-16.33) | 12.93 (9.70-17.25) |
| **2019 under-5 severe wasting prevalence[7]** | 1.13 (0.67-1.77) | 0.85 (0.38-1.65) |
| **2022 Corruption Perceptions Index[8]** | 26 / 100 | 17 / 100 |
| **2022 GDP per capita[9]** | $1588.5 | $1748.3 |
| **2015 Healthcare Access and Quality Index[10]** | 44.4 (35.0-53.3) | 38.5 (33.7-43.5) |
| **Gini Index[11]** | 46.6 | 41.1 |
| **2018 Wellcome Global Monitor: Trust in neighbors[12]** | 44% | 67% |
| **2018 Wellcome Global Monitor: Trust in government[12]** | 51% | 46% |
| **2018 Wellcome Global Monitor: Trust in scientists[12]** | 49% | 62% |
| **Vaccine Confidence Project: Vaccines are important[13]** | 82.98% (71.83-91.84) | 91.18% (82.80-97.49) |
| **Vaccine Confidence Project: Vaccines are safe[13]** | 56.50% (42.46-72.76) | 62.33% (43.10-77.75) |
| **Vaccine Confidence Project: Vaccines are effective[13]** | 63.24% (45.55-83.46) | 71.80% (51.30-88.96) |
| **2020 Average precipitation in depth (mm per year)[14]** | 1,604 | 1,440 |
| **Population density (people per sq. km of land area)[15]** | 58 | 415 |
| **Percent of population exposed to high flood risk[16]** | 19.1 | 17.5 |

MCV= Measles containing vaccine; ORT = Oral rehydration therapy; GDP = Gross domestic product

Improved water access includes piped water and other improved sources; improved sanitation access includes sewer and septic as well as other improved sources. For the Corruption Perceptions Index, a lower score indicates more corruption. Gini data were most recently available for 2014 in Cameroon and 2012 in Haiti. Wellcome Global Monitor trust data use “a lot” or “some” trust as survey answers indicating trust. Vaccine confidence data were pulled from modeled estimates and used the midpoint of 2018 as the timestamp; importance, safety, and efficacy were defined as the portion of respondents who replied “strongly agree”.

Table 2: Number and percent of vaccine effectiveness observations outside 0% or 100%, Cameroon and Haiti

| | Cameroon (n=196 data points) | Haiti (n=302 data points) |
|---|---|---|
| **Upper 95% CI estimates** | 3 (1.5%) | 2 (0.7%) |
| **Lower 95% CI estimates** | 24 (12.2%) | 3 (1.0%) |
| **Median estimates** | 5 (2.6%) | 2 (0.7%) |

We noted a small percentage of estimates that fell outside of the 0 to 100% boundary. This is likely a result of using static estimates of vaccine coverage over the entire duration of the vaccine study period rather than dynamic estimates, which were unavailable for OCV1 campaigns in either country. However, in our study, we were most interested in understanding general patterns of vaccine effectiveness over time and interpreting the median estimates and whether credible intervals overlapped. As such, a small percentage of results out-of-bounds—especially in the credible interval—do not drastically alter our interpretation of the results. In clinically-motivated vaccine effectiveness studies where precision is more important, time-varying vaccine coverage should be considered for similar analysis.

### Section 2: Sensitivity Analyses

In order to understand the impact of our assumptions on our findings, we changed our assumptions in a number of ways, each independently while holding all other variables constant.

#### *Artificially changing OCV coverage to understand impact on vaccine effectiveness, Cameroon and Haiti*

For our main analysis, we used reported coverage figures for OCV1 campaigns in Cameroon and Haiti. Given that these estimates are highly localized to the targeted locality and do not reflect the coverage status in the whole country, we explored how changing coverage from 60% OCV1 coverage to 90% OCV1 coverage would impact estimated OCV1 effectiveness in both Cameroon and Haiti. When assuming coverage was at 60% rather than the reported 85.50% and 76.00% in Cameroon and Haiti, respectively, vaccine effectiveness was 76.48% (15.24-122.29%) and 95.23% (66.66-108.07%). In contrast, assuming coverage of 90% revealed vaccine effectiveness estimates of 50.99% (10.16-81.52%) in Cameroon and 63.49% (44.44-72.05%) in Haiti.

<u>Figure 1: Impact of assumed OCV1 coverage on vaccine effectiveness</u>

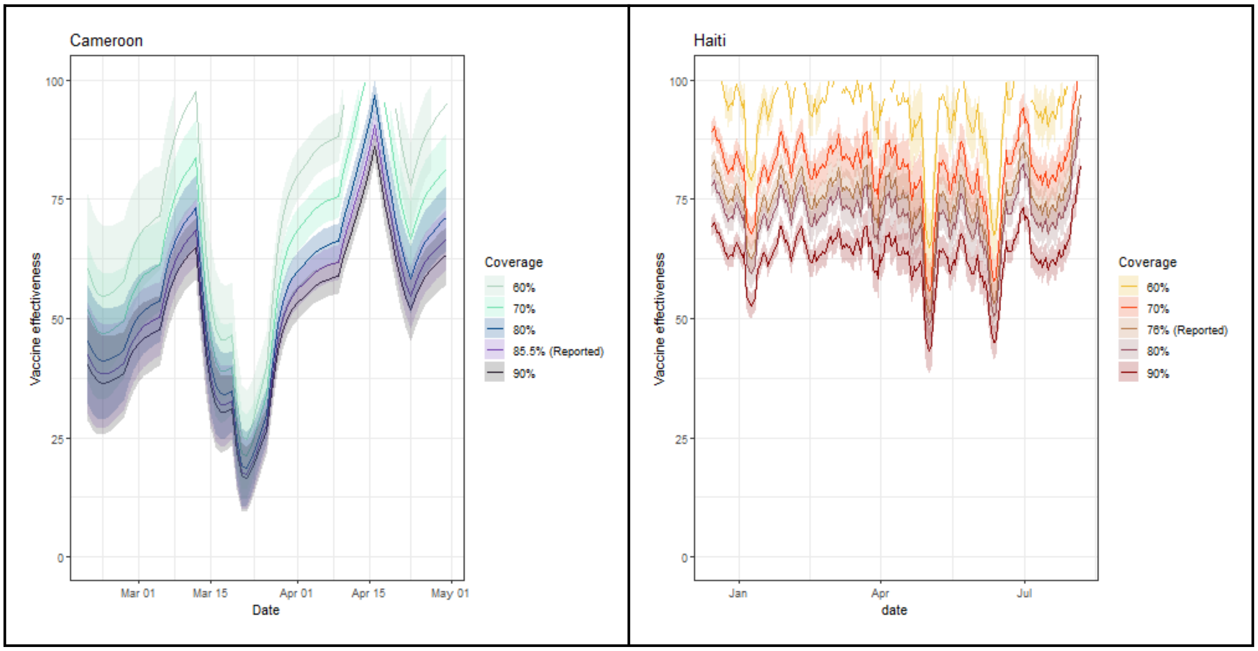


#### *Exploring alternate $R_t$ windows in EpiEstim, Cameroon and Haiti*

While we chose a $R_t$ window of one-week to optimize the relationship between too much statistical noise with smaller windows and too much smoothing with larger windows, we explored additional time windows: four days and two weeks. While the shorter window had higher initial $R_0$ values and the longer window had fewer days under $R_t$=1, qualitatively the patterns across all three windows analyzed were relatively consistent.

Figure 2: Impact of length of time window on $R_t$

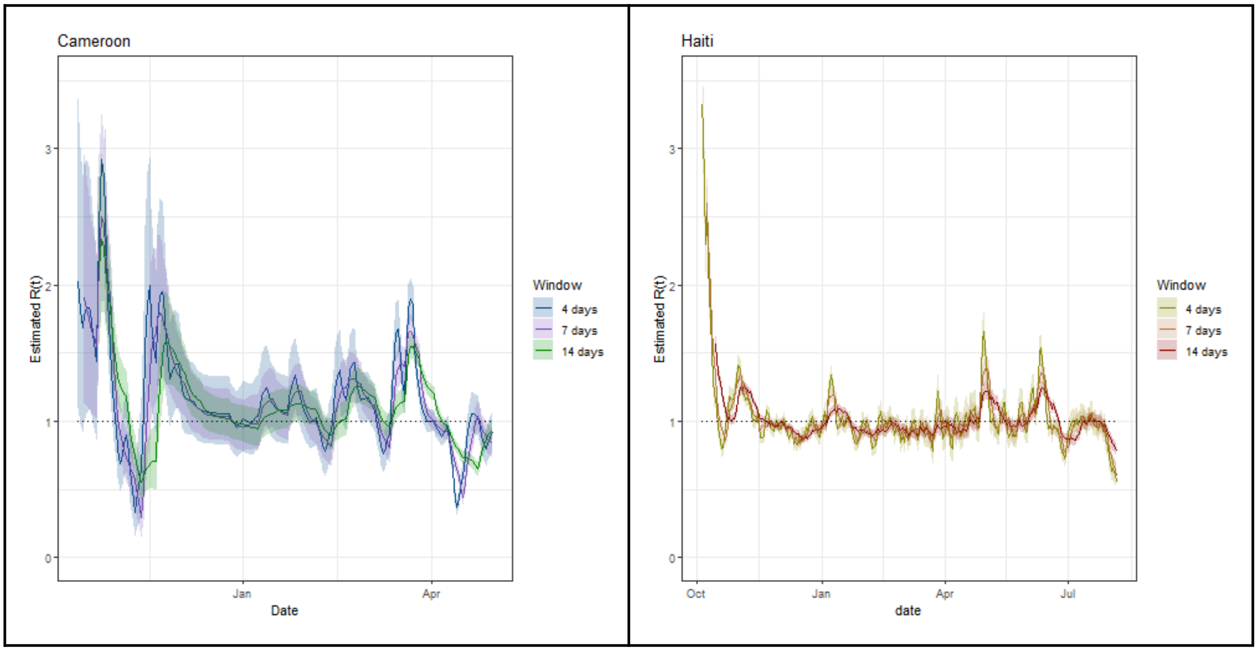


#### *Exploring alternate SI parameters in EpiEstim, Cameroon and Haiti*

For our SI, we used a gamma distribution with a mean of 4 days and a SD of 3 derived from previous studies of cholera among household contacts,[17] historic investigations of cholera,[18] and used among previous EpiEstim analyses of cholera.[19,20] However, studies of cholera in humanitarian and crisis settings used a slightly higher and wider distribution with a mean of 5 and a standard deviation of 8.[21–23] We modified our analysis to use this wider distribution and qualitatively found similar results between the two distributions, with the narrower SI having slightly higher peaks and lower troughs, but generally similar patterns.

Figure 3: Impact of changing the SI on $R_t$

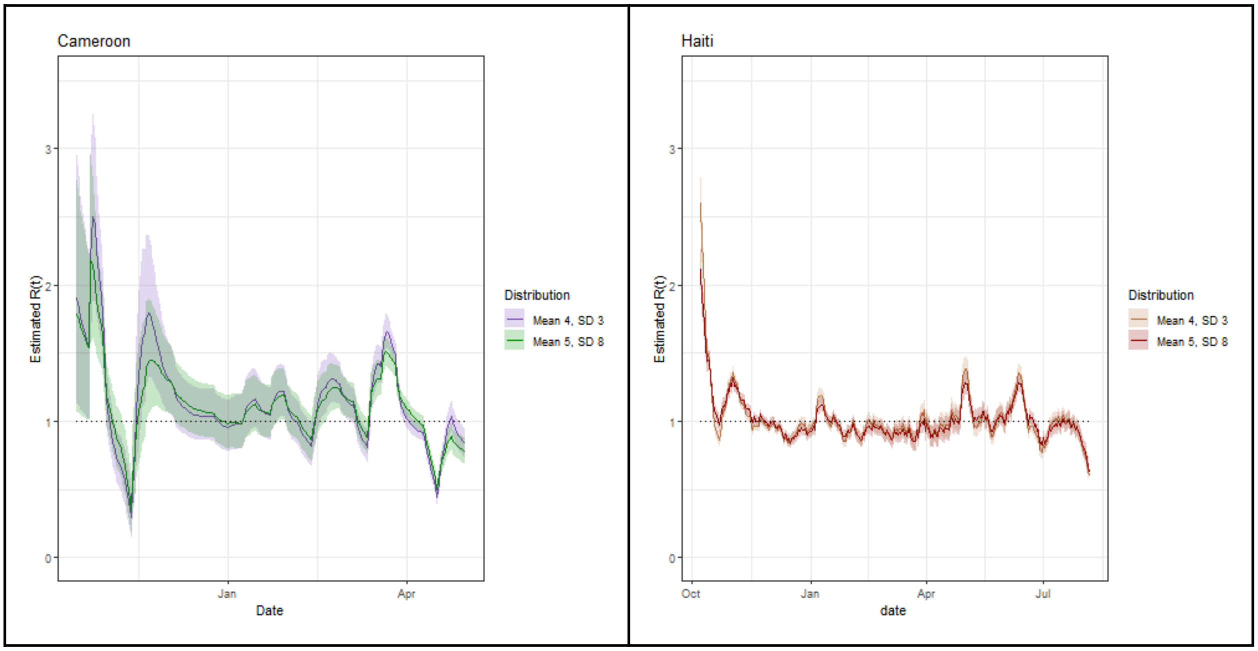


#### *Exploring alternate values of $R_0$ on vaccine effectiveness, Cameroon and Haiti*

In our main analysis, we used averages of the published regional and local data for $R_0$ for our two countries of interest. For our sensitivity analysis, we considered the minimum $R_0$ published in the literature (1.10 Cameroon / 1.06 Haiti) and maximum published $R_0$ (3.5 Cameroon / 3.72 Haiti) in each location. In contrast to the changes to the window and the SI which had few qualitative impacts on our findings, changing $R_0$ drastically altered the vaccine effectiveness estimates. Using the minimum $R_0$ value in each Cameroon in Haiti resulted in median estimates of vaccine effectiveness of 19.41% (6.40-61.39%) and 12.22% (1.61-35.78%), respectively, while using the maximum $R_0$ value in each country resulted in average vaccine effectiveness of 83.08% (62.29-97.69%) and 96.97% (83.88-99.70%) for both Cameroon and Haiti. In addition, in both countries, the relative relationship overtime remained consistent despite the changing magnitude of the estimates. It is important to note that these minima and maxima represent estimates at extreme ends of the credible intervals of the distribution of $R_0$ and therefore are meant to be considered as the extremes in the analysis of the vaccine effectiveness estimates rather than those that were most probable.[20,24] Notably, such extreme choices of $R_0$ also resulted in a larger proportion of estimates falling outside of the bounded range of 0 to 100%, suggesting that while these are within the realm of possible choices for $R_0$, they are unlikely to be the realized values of $R_0$ during the outbreaks in Cameroon and Haiti.

Figure 4: Impact of changing $R_0$ on OCV1 effectiveness

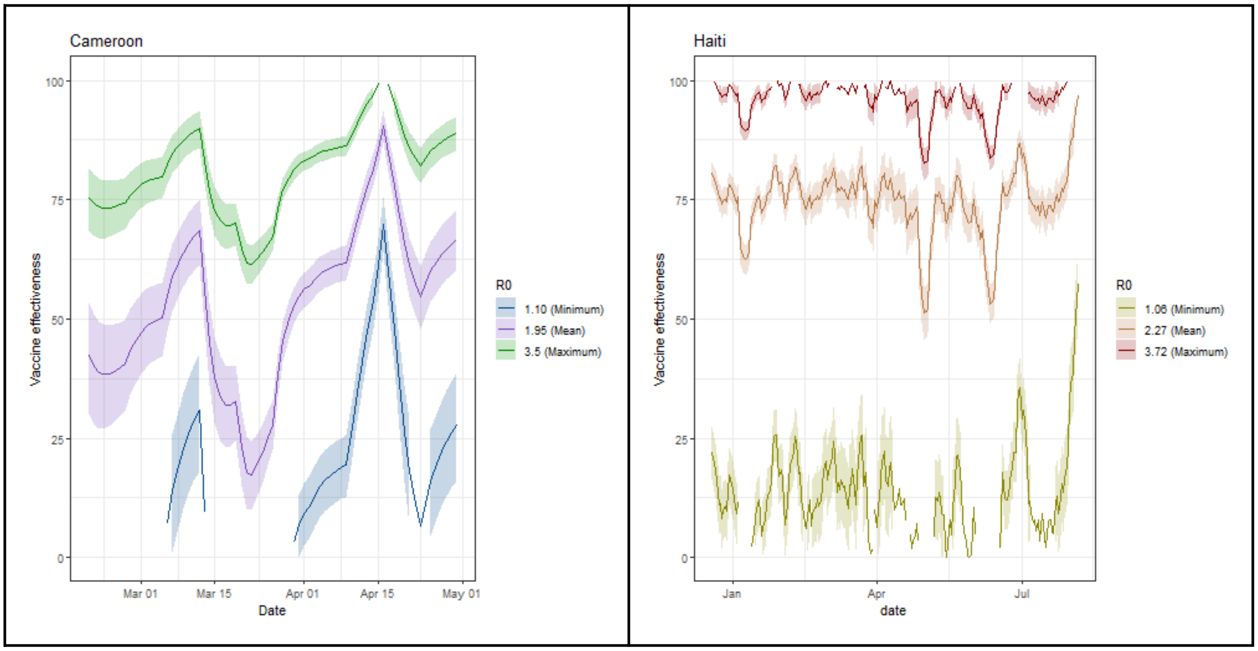


#### *Exploring the vaccine effectiveness in Ouest department, Haiti*

In Haiti, where the OCV1 campaign was highly localized to the Ouest and Centre departments, we also conducted a sensitivity analysis using daily cholera data published by department by Haiti's Ministry of Health (MSPP)[25] to understand how localized data influence our results. We consider the vaccine effectiveness in the Ouest department of Haiti using digitized daily cholera data (digitized using WebPlotDigitizer[26]) from October 1, 2022 to August 5, 2023, using reported OCV1 coverage at 69.90%.[27,28,25] This analysis suggests a median vaccine effectiveness of 83.11% (95% UI: 36.20-97.26%) over the entire vaccination period after December 19, 2022, almost 10% higher than the point estimate at the national level; however, the credible interval overlapped with the national level analysis, and thus this difference was not considered statistically significant.

Figure 5: Focusing on Ouest department for vaccine effectiveness in Haiti

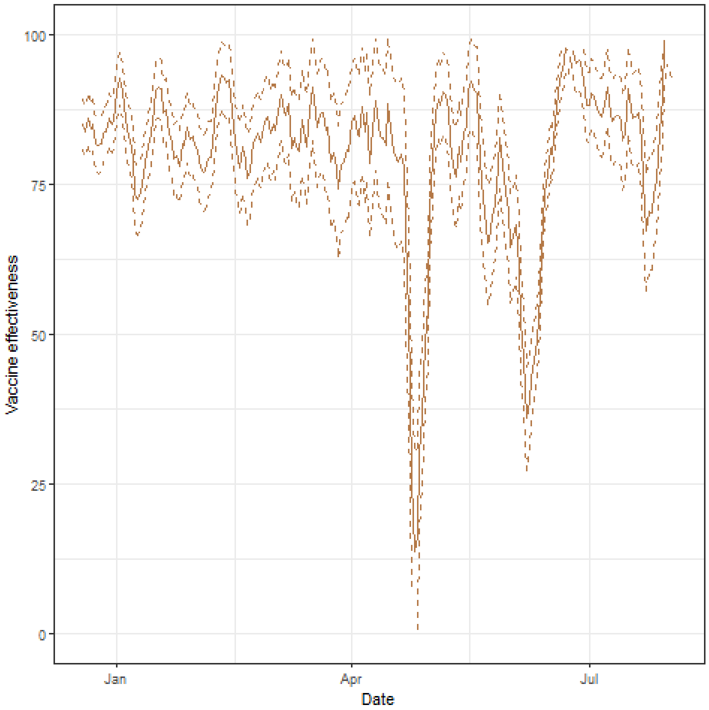
